# Predictors of mortality among low birth weight neonates after hospital discharge in a low-resource setting: A case study in Uganda

**DOI:** 10.1101/2023.07.01.23292109

**Authors:** Ronald Nsubuga, Joseph Rujumba, Saleh Nyende, Stevens Kisaka, Richard Idro, Jolly Nankunda

## Abstract

**Background:** Most neonatal deaths occur among low birth weight infants. However, in resource-limited settings, these infants are commonly discharged early which further exposes them to mortality. Previous studies on morbidity and mortality among low birth weight infants after early discharge mainly focused on very low birth weight infants, and none described post-discharge neonatal mortality. This study aimed to determine the proportion and predictors of mortality among low birth weight neonates discharged from the Special Care Baby Unit at Mulago National Referral Hospital in Uganda.

**Methods:** This was a prospective cohort study of 220 low birth weight neonates discharged from the Special Care Baby Unit at Mulago National Referral Hospital. These were followed up to 28 completed days of life, or death, whichever occurred first. Proportions were used to express mortality. To determine the predictors of mortality, Cox hazards regression was performed.

**Results:** Of the 220 enrolled participants, 216 (98.1%) completed the follow-up. The mean gestational age of study participants was 34 ±3 weeks. The median weight at discharge was 1,650g (IQR: 1,315g -1,922g) and 46.1% were small for gestational age. During follow-up, 14/216 (6.5%) of neonates died. Mortality was highest (7/34, 20.6%) among neonates with discharge weights less than 1,200g. The causes of death included presumed neonatal sepsis (10/14, 71.4%), suspected aspiration pneumonia (2/14, 14.3%), and suspected cot death (2/14, 14.3%). The median time to death after discharge was 11 days (range 3-16 days). The predictors of mortality were a discharge weight of less than 1,200g (adj HR: 23.47, *p* <0.001), a 5-minute Apgar score of less than 7 (adj HR: 4.25, *p* = 0.016), and a diagnosis of neonatal sepsis during admission (adj HR: 7.93, *p =* 0.009).

**Conclusion:** Post-discharge mortality among low birth weight neonates at Mulago National Referral Hospital is high. A discharge weight of less than 1,200g may be considered unsafe among neonates. Caregiver education about neonatal danger signs, and measures to prevent sepsis, aspiration, and cot death should be emphasized before discharge and during follow-up visits.

## Introduction

Low birth weight is defined as weight at birth of less than 2500g(1) Small for gestational age (SGA) is defined as weight for gestational age below the 10^th^ percentile(2)

Worldwide, an estimated 20 million infants are born with low birth weight (LBW) annually. Most of these births (96.5%) occur in low and middle-income countries (LMICs)(3). Low birth weight contributes over 80% of all neonatal deaths worldwide(4). Low birth weight neonates are at increased odds of death compared to their heavier counterparts(5) and LBW is considered the single most important predictor of infant mortality (6).

The World Health Organization (WHO) developed discharge criteria for LBW infants(7). However, this has been difficult to adhere to in resource-limited settings like Uganda with an over-stretched healthcare system and an estimated 10% of births have LBW(8). In Uganda, many neonatal units lack proper Kangaroo Mother Care (KMC) facilities(9,10), yet this is known to improve the survival of LBW infants (11).

Previous studies have assessed the morbidity and mortality of LBW infants after early discharge. Nonetheless, these mainly focused on Very Low Birth Weight (VLBW) infants and without the description of post-discharge neonatal mortality (12–14). Post-discharge outcomes of LBW neonates are largely unknown.

In our setting, LBW infants are discharged from the Special Care Baby Unit (SCBU) once stable (off antibiotics and oxygen, with stable body temperature in a cot). In addition, they should be able to tolerate 10ml per feed of expressed breastmilk by cup or tube, regardless of body weight or weight growth velocity. These babies are followed up weekly until they attain a body weight of 2500g. This practice may serve to reduce pressure on available limited resources, but its safety has not been assessed.

This study aimed at determining the proportion of LBW neonates that die after discharge from the SCBU, and the predictors of mortality. The study findings may be used to design targeted strategies to reduce mortality among low birth weight neonates after hospital discharge.

## Methods and materials

### Study setting

This was a prospective cohort study conducted at the SCBU of Mulago National Referral Hospital from 8^th^ November 2018 to 28^th^ January 2019. This SCBU was a level II neonatal unit. On average 4,600 babies are admitted to the unit annually, and 50% of these are low birth weight neonates.

The SCBU had an official bed capacity of 50 with 13 baby cots, 22 incubators, 11 radiant warmers, and 4 beds, but it often houses over 100 babies. It had term and preterm baby sections but with no KMC wing. The unit was run by 4 Paediatricians, 4-6 Paediatrics residents, 2 intern doctors on a rotational basis, and 18 nurses (working in 3 shifts a day).

The care offered included the provision of warmth (incubators and radiant warmers), phototherapy, and bubble Continuous Positive Airway Pressure (CPAP). Admitted neonates may also receive intravenous fluids (two hourly boluses), intravenous antibiotics, and nasogastric feeding. Vital signs were intermittently assessed and caregivers feed their babies every two hours under the supervision of the nursing team.

Low birth weight infants are discharged from the SCBU once they are deemed physiologically stable (normal axillary temperature, off oxygen, off intravenous fluids, and apnea-free), tolerating at least 10ml per enteral feed, and gaining weight. The caregiver must demonstrate knowledge and competence in the provision of care to the infant before discharge. On average, 138 LBW neonates are discharged monthly.

After discharge, LBW infants were followed up in the clinic every week until a body weight of 2500g is attained. During clinic visits, growth monitoring, drug refills (Iron and multivitamins), nasogastric tube replacement, and adjustment of feeds were done. Unwell infants were identified and readmitted.

### Study population

All low birth weight neonates discharged from the SCBU whose parents provided written consent were included in the study. Those who were neither reachable physically nor by telephone were excluded from the study.

### Sample size

The sample size was estimated using an OpenEpi formula for frequency in a population, (15).

Sample size ***n* = [DEFF*Np(1-p)]/ [(d**^2^**/Z**^2^_1-α/2_ **\*(N-1)+p*(1-p)]**

Where;

DEFF was the sampling design effect = 1 since we were not doing multi-stage sampling. N is the population size (for finite population correction factor) = 135*3= 405

p is the expected proportion of LBW neonates that die after discharge from SCU= 50% since there were no published studies in Africa about post-discharge neonatal mortality among LBW infants.

d is the acceptable absolute error = 5%.

z is the standard normal distribution value corresponding to 95% Confidence Level. Sample size (n) for 95% Confidence Level = 198 participants.

Considering a 10% loss to follow-up rate, the final sample size (n) was 220.

### Sampling procedure

Study participants were recruited consecutively until the sample size was achieved.

### Data collection procedure

All neonates with birth weight <2500g were identified by a research assistant at discharge. Their parents were then introduced to the study. For neonates who met the inclusion criteria, the research assistant obtained written informed consent from the caregiver/ guardian. Recruitment was conducted daily and consecutively until the desired sample size was achieved.

Recruited neonates received a study identification number and their details were captured in a logbook. Each parent provided 3 phone contacts for purposes of follow-up. Phone contacts of the principal investigator (PI) and research assistants were shared with parents for them to call in case of any concerns.

Data were captured at discharge using a pre-tested checklist and questionnaire. At every clinic visit, a follow-up chart was filled. Study participants were followed up until 28 completed days of life. Parents of study participants were contacted by telephone once a week and significant events (death, illness) were registered. The possible cause of death was established by the Principal investigator using the 2016 World Health Organization verbal autopsy instrument (16) Those who missed their scheduled follow-up visits were contacted by phone and requested to bring the neonate for review on the next clinic day. A neonate who missed two consecutive clinic visits and whose parents were not reachable by phone throughout the study period was considered lost to follow-up.

### Data collection tools

The data checklist had the following sections: Maternal factors, and neonatal factors.

The questionnaire had the following sections: Maternal, delivery, and post-delivery factors, neonatal factors, and socioeconomic factors including a multidimensional social support scale.

The follow-up chart had the following sections: post-discharge follow-up body weight record in grams, readmission details, and mortality details where applicable

### Data management and analysis

Captured data were entered into Epidata version 3.1, checked for accuracy and completeness, and exported to STATA Version 14 for analysis. Continuous variables were summarized using the mean and standard deviation, and median and interquartile ranges for skewed data. Categorical variables were summarized using frequencies and proportions.

The proportion of LBW neonates that died after discharge from the SCU was expressed as a percentage. The numerator was the number of babies that died within the first 28 days of life and the denominator was the total number of LBW neonates recruited into the study. Chi-square and Fisher’s exact tests were used to compare the proportion of LBW neonates that died after discharge from the SCU by their baseline characteristics.

Student-t and Wilcoxon rank sum tests were used to compare means and median respectively of the continuous variables of participants that died and those that were alive after 28 completed days of life. To determine the predictors of mortality, relative death rates (Hazard Ratios) were obtained using simple Cox proportional hazards regression.

Variables that achieved a significance level of ≤0.05 were then included in a multivariable Cox proportional hazards model. Backward elimination was used to build a model of best fit. The significance level was set at ≤0.05 and a confidence interval of 95% was used. Kaplan Meier curves were used to compare survival across gender, gestation age at birth, and birth weight.

### Ethical approval and consent to participate

This study protocol was subjected to approval by the Makerere University School of Medicine Research Ethics Committee. The investigators ensured that caregivers (parents or guardians) of study participants were given full and adequate oral and written information about the nature, purpose, possible risks, and benefits of the study. They were given adequate opportunities to ask questions and allowed time to consider the information provided voluntarily. Written consent was obtained from caregivers before conducting this study. Participants in this database were identified by their unique enrollment numbers. Only the PI had access to the participant identification list, which included their unique codes, full names, phone numbers, and address.

## Results

Of the 243 LBW neonates identified for potential recruitment into the study, 5 did not consent to the study, and 18 did not have phone contact. Therefore, a total of 220 (representing 90.5%) neonates were enrolled in the study. Of these, 216 completed follow-up **(Figure 1)**. The mean gestational age at birth was 34 ±3 weeks, and the median weight at discharge was 1650g (IQR 1315-1922 grams). All four neonates who were lost to follow-up had discharge weight >2000g.

The median length of hospital stay of study participants was 5 days (IQR 3-8 days). The majority of the study participants (134/216, 62%) were fully breastfeeding, and the rest were breastfeeding and receiving top-up feeds through a nasogastric tube. Five study participants had confirmed neonatal sepsis during admission, and they had completed treatment before discharge (**Table 1** shows the baseline characteristics of the study participants).

**Table 1.**
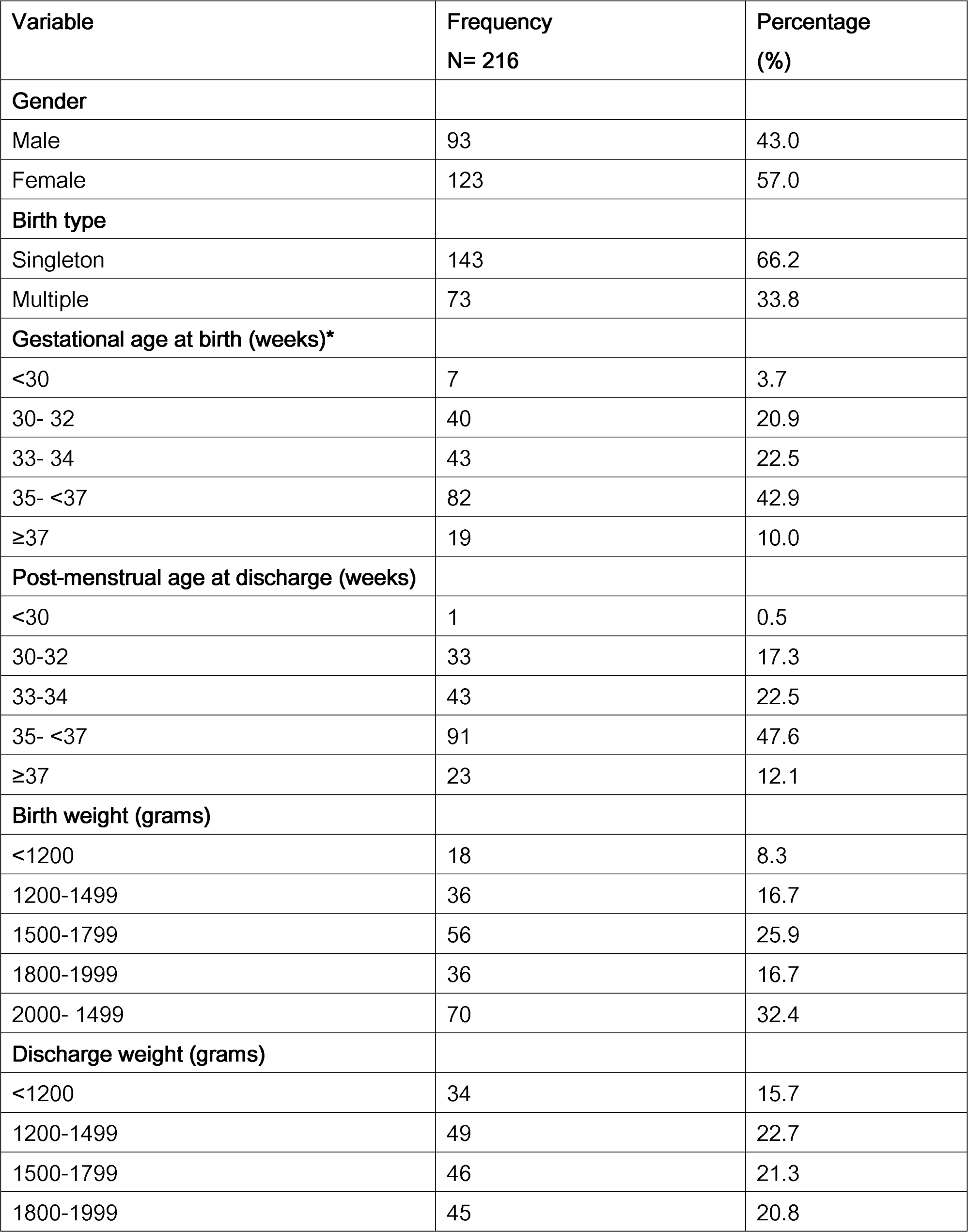
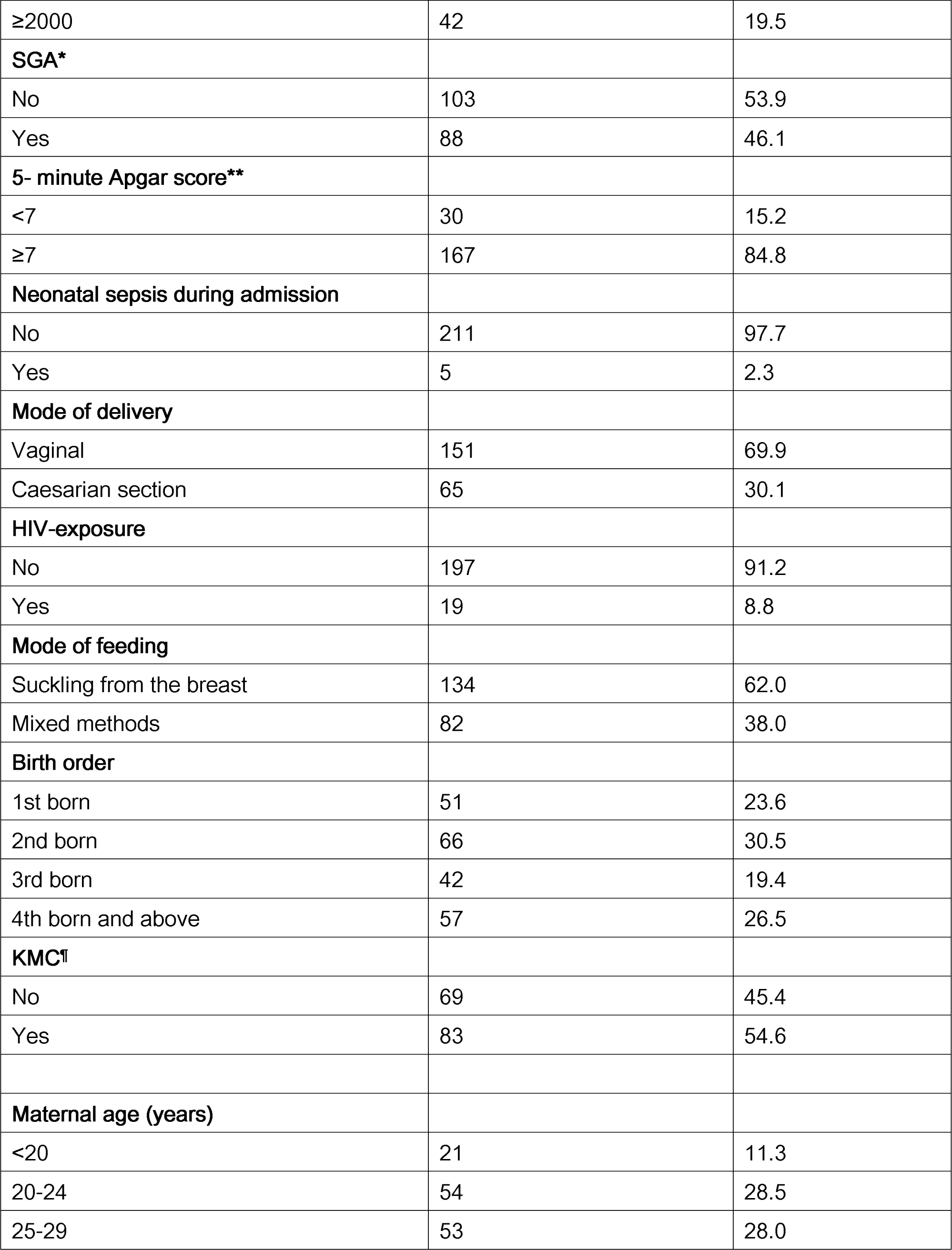
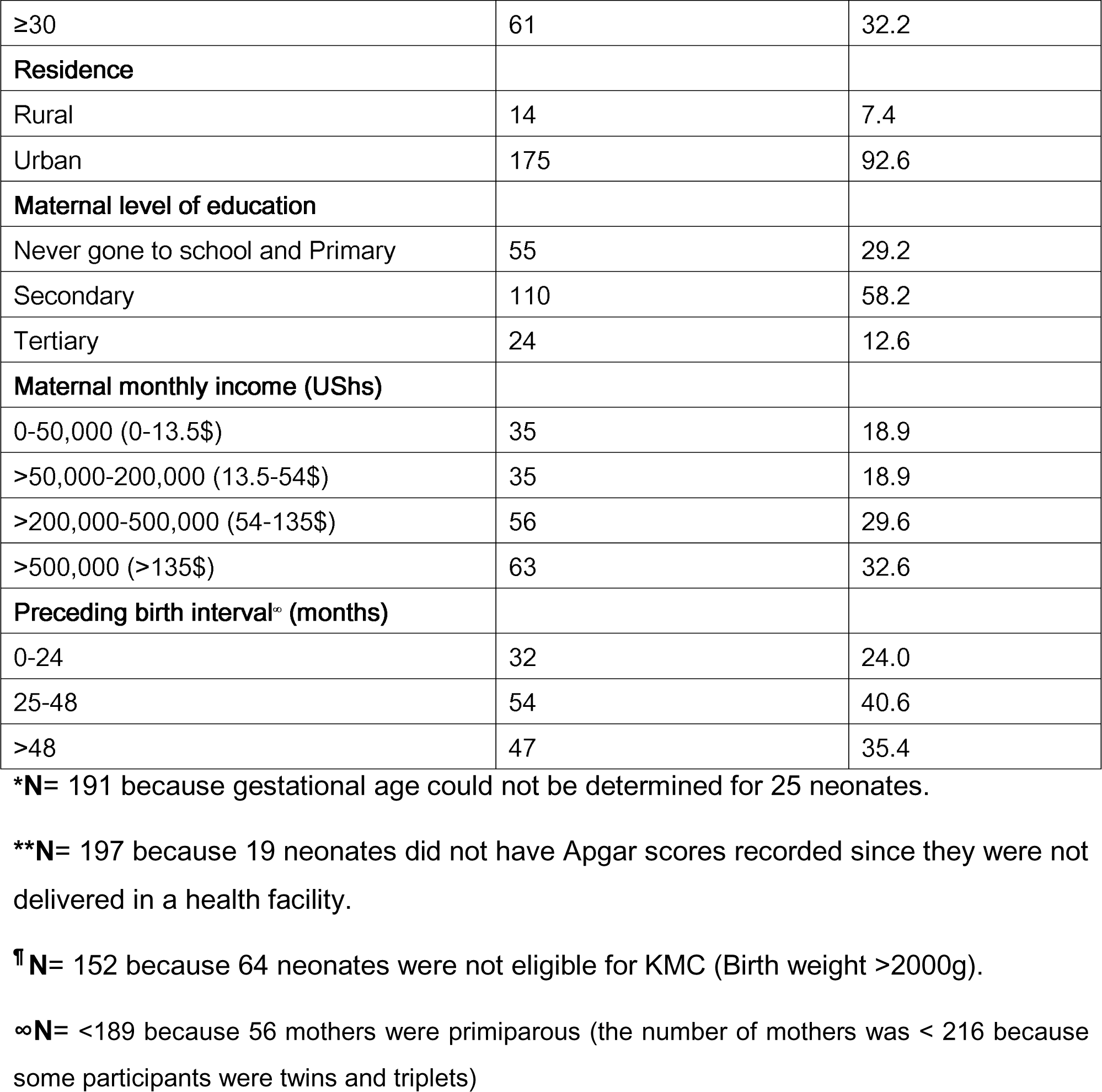
Neonatal and maternal baseline characteristics.

During follow-up, 14/216 (6.4%) neonates died. Of these, 8 occurred from home, 5 occurred at a health facility, and 1 occurred on the way to the hospital. The circumstances surrounding each death were established by contacting the primary caregiver by phone. The possible causes of death on verbal autopsy included neonatal sepsis (71.4%), aspiration pneumonia (14.3%), and cot death (14.3%). The median time to death after discharge was 11 days (range 3-16 days).

The proportion of neonates that died was highest among those with discharge weight <1200g (20.6%) and gestational age at birth <30 weeks (28.6%).

Figure 2 shows that the overall survival probability was 93.5% and the greatest drop in survival probability occurred in the first week after discharge. The median survival time had not been reached by the time the study closed meaning that survival was sustained above 50% throughout the study period. Survival was compared according to birth weight. Figure 3 shows that survival probability was highest among study participants with birth weight >2,000g. Notably, survival dropped with reducing birth weight.

The neonates who died had a lower median body weight at birth, discharge, and first follow-up visit, compared to those who survived. In both groups, the median discharge weight was lower than the birth weight. Among those who died, the median body weight at the first follow-up visit was lower than the median weight at discharge as shown in Figure 4.

Predictors of mortality were assessed at bivariate and multivariate levels of analysis.

At bivariate analysis of neonatal and maternal characteristics, birth weight, discharge weight, 5-minute Apgar score, post-menstrual age at discharge, and neonatal sepsis at admission were statistically significant (Table 2).

**Table 2.**
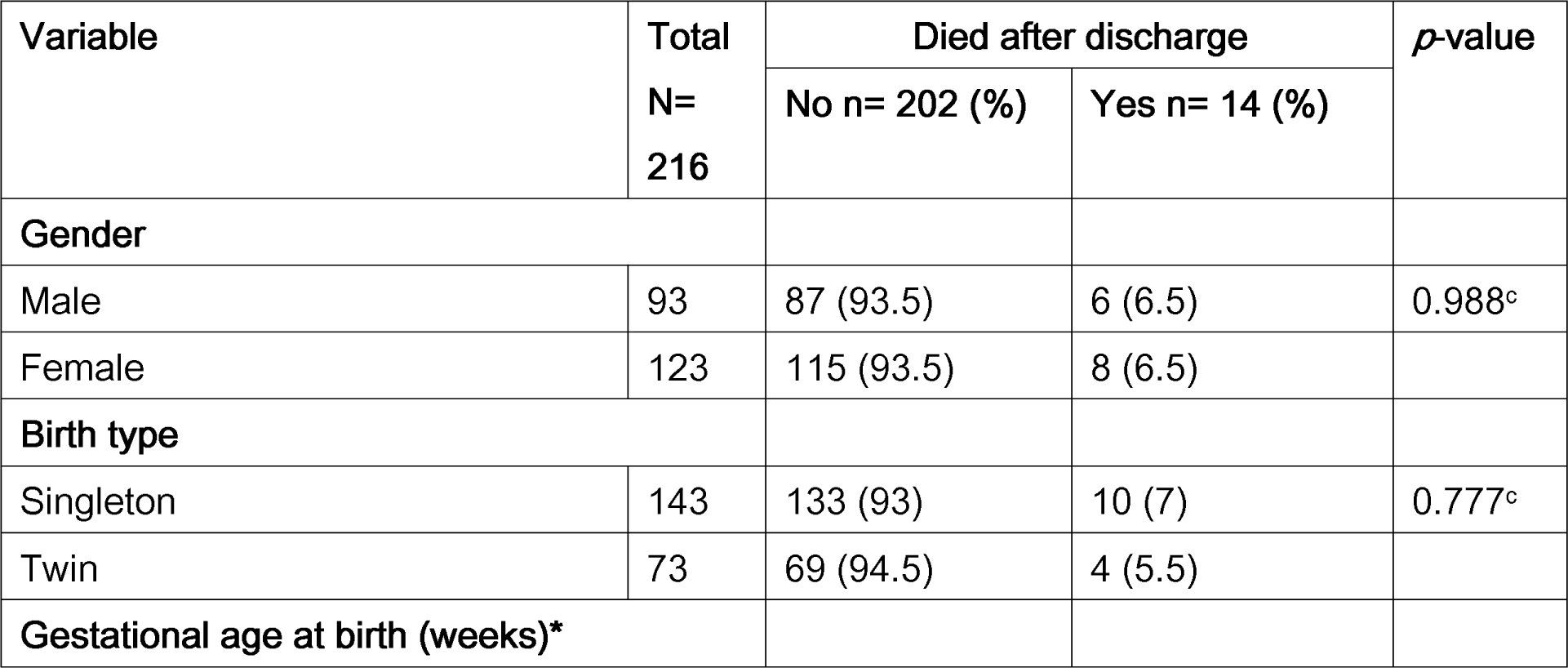
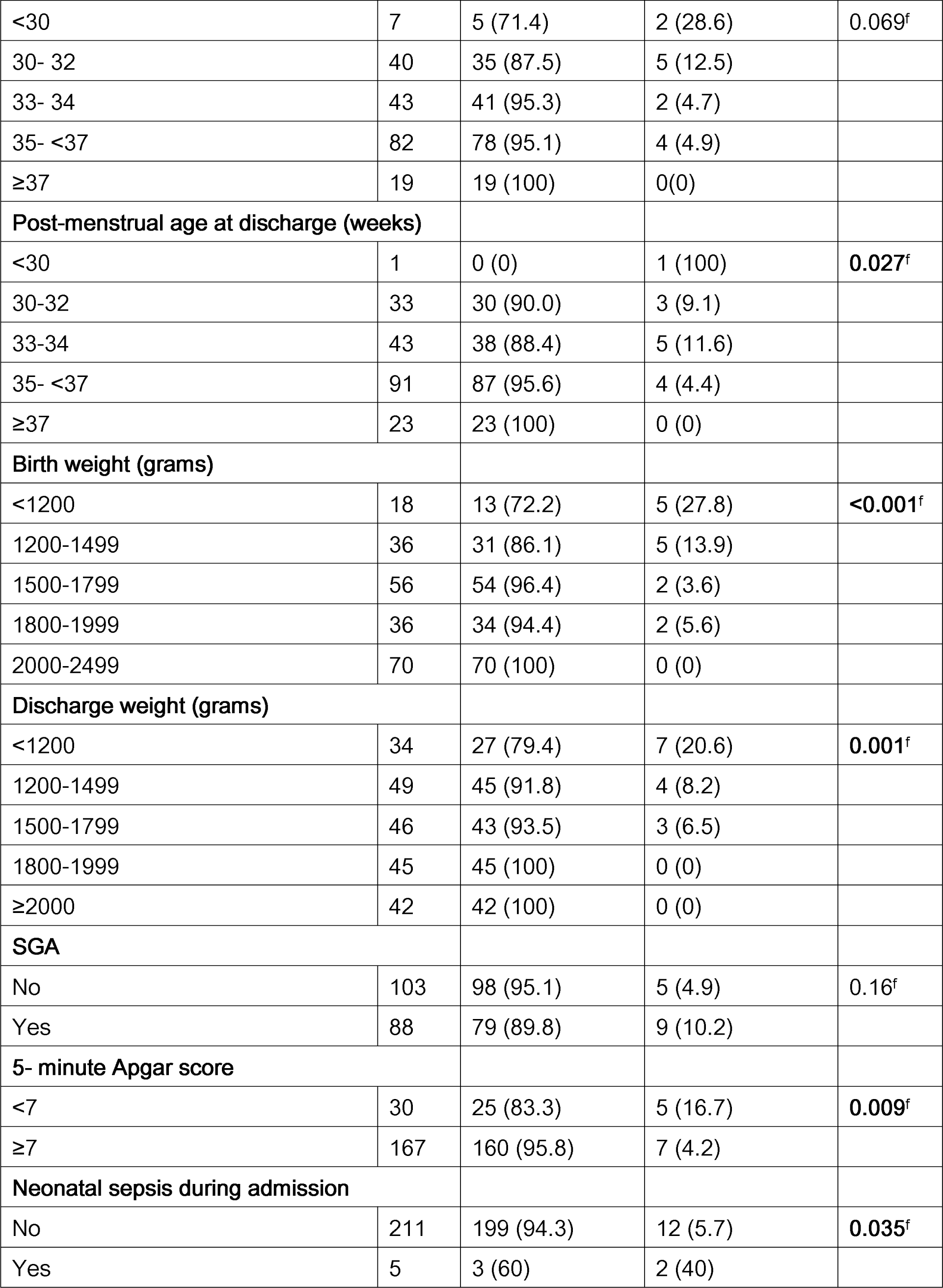
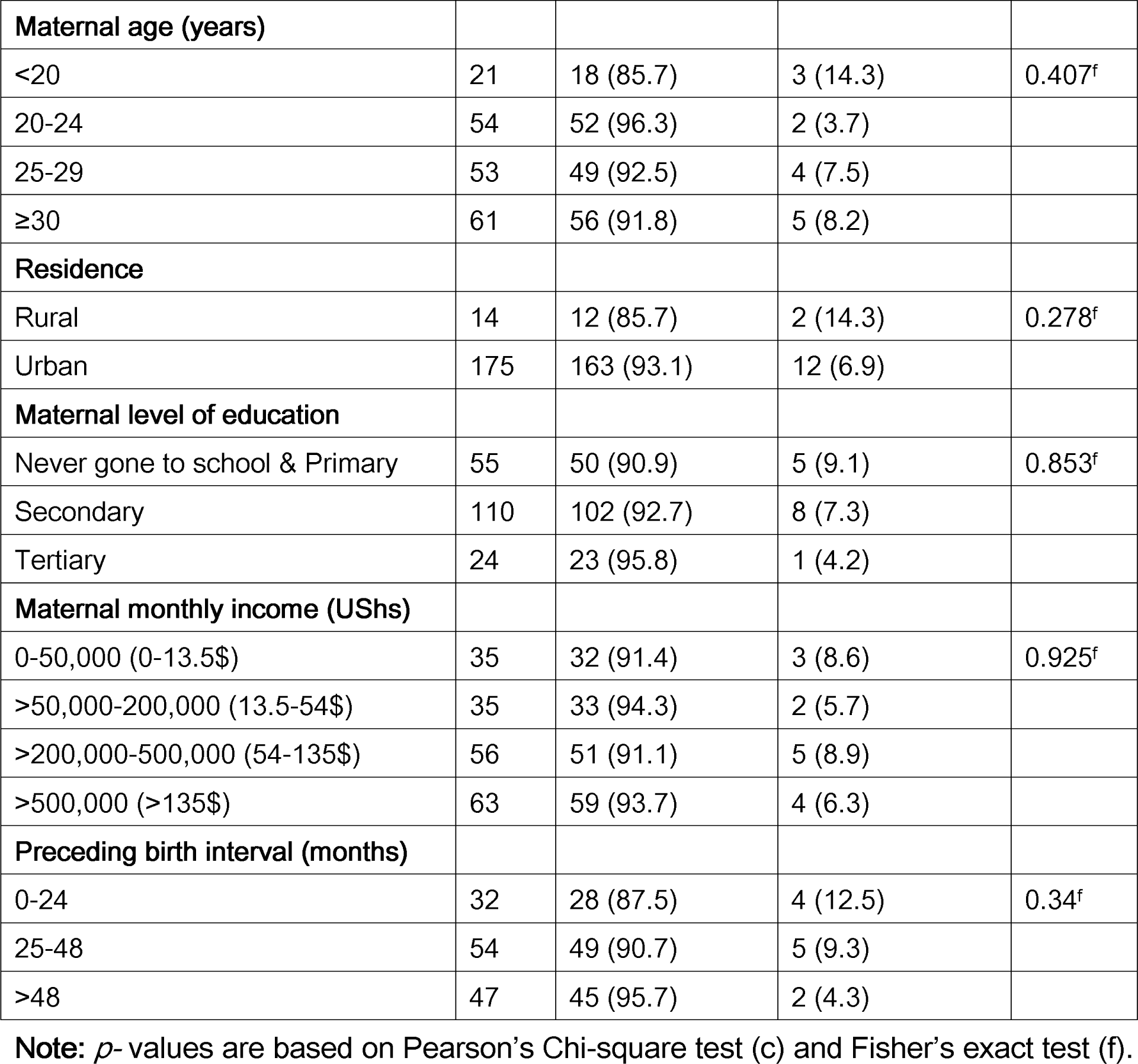
Bivariate analysis of neonatal and maternal characteristics, and post-discharge mortality.

At multivariate analysis, discharge weight <1200g was associated with a 23.5-fold increase in the risk of death compared to babies of discharge weight greater than 1,500g (p<0.001). The neonates who posted a 5-minute Apgar of <7 were 4.42 times more likely to die compared to those with a 5-minute Apgar of 7 and above (p=0.016). Similarly, having a diagnosis of neonatal sepsis at admission was significantly associated with mortality compared with those who did not have such a diagnosis (adj HR = 7.93, p = 0.009). Notably, none of the maternal sociodemographic characteristics was significantly associated with mortality. The details of the unadjusted and adjusted hazard ratios are shown in Table 3.

**Table 3.**
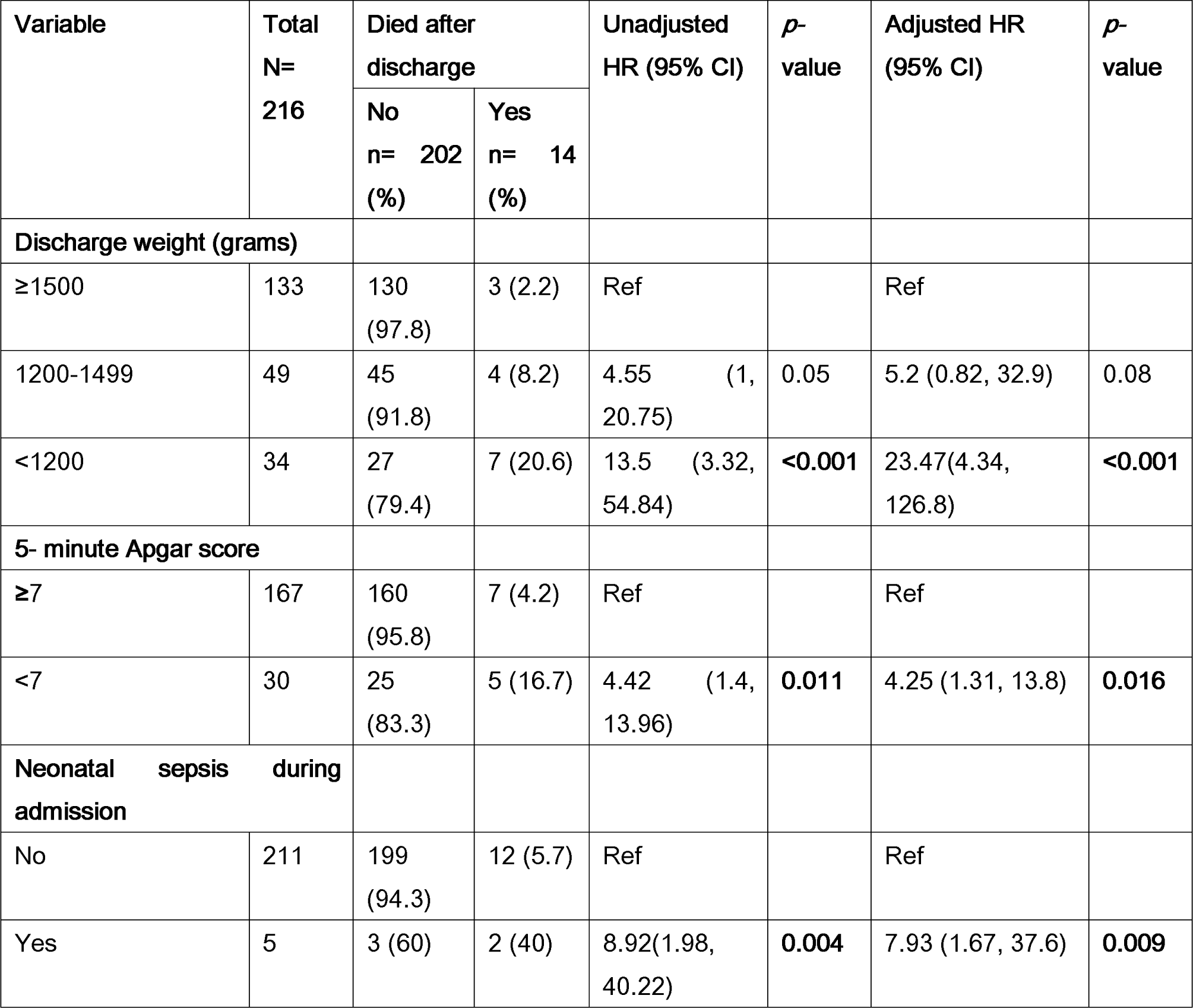
Multiple Cox regression for predictors of mortality among the 216 low birth weight neonates after discharge from the SCU.

| Variable | Total<br>N=<br>216 | Died after<br>discharge |  | Unadjusted<br>HR (95% CI) | *p*-<br>value | Adjusted HR<br>(95% CI) | *p*-<br>value |
| --- | --- | --- | --- | --- | --- | --- | --- |
|  |  | No<br>n= 202<br>(%) | Yes<br>n= 14<br>(%) |  |  |  |  |
| Discharge weight (grams) |  |  |  |  |  |  |  |
| ≥1500 | 133 | 130<br>(97.8) | 3 (2.2) | Ref |  | Ref |  |
| 1200-1499 | 49 | 45<br>(91.8) | 4 (8.2) | 4.55 (1,<br>20.75) | 0.05 | 5.2 (0.82, 32.9) | 0.08 |
| <1200 | 34 | 27<br>(79.4) | 7 (20.6) | 13.5 (3.32,<br>54.84) | <0.001 | 23.47(4.34,<br>126.8) | <0.001 |
| 5- minute Apgar score |  |  |  |  |  |  |  |
| ≥7 | 167 | 160<br>(95.8) | 7 (4.2) | Ref |  | Ref |  |
| <7 | 30 | 25<br>(83.3) | 5 (16.7) | 4.42 (1.4,<br>13.96) | 0.011 | 4.25 (1.31, 13.8) | 0.016 |
| Neonatal sepsis during admission |  |  |  |  |  |  |  |
| No | 211 | 199<br>(94.3) | 12 (5.7) | Ref |  | Ref |  |
| Yes | 5 | 3 (60) | 2 (40) | 8.92(1.98,<br>40.22) | 0.004 | 7.93 (1.67, 37.6) | 0.009 |

## Discussion

In this study, we observed a post-discharge neonatal mortality of 6.5%. Mortality was highest among neonates delivered before 30 weeks of gestation (28.6%), those with birth weight <1500g (18.5%), and discharge weight <1200g (20.6%). Most of the deaths occurred at home and were likely due to possible neonatal sepsis.

The post-discharge mortality we observed at the referral hospital was lower than what was previously reported in two other similar settings. In Bangladesh, a study by Yasmin et al, 2001 found a mortality of 13.3% (5), while in Malawi, Blencove et al, 2009 found a mortality of 12.4% (17). The discrepancies may be explained by advances in neonatal care over time. The level of mortality was however comparable to what Vazirinejad et al, 2012 found (5.4%) in Iran(18), and what Kibona et al found (5%) in Tanzania (19).

In all these studies, however, mortality was similarly higher among neonates with birth weights <1500g and those with gestational age at birth <30 weeks (20–23). The lower mortality observed in our study and Tanzania may be due to advances in neonatal care over the last 10 years since the other two studies were carried out. The higher mortality among those with birth weight <1500g, gestational age at birth <30 weeks, or discharge weight <1200g may be explained by immature oromotor coordination, poor homeostatic control among neonates with lower birth weight and lower gestational age at birth(24), and increased risk of sepsis in the same age group.

In our study, 3 in 5 LBW neonates who died had possible neonatal sepsis. This is comparable to the findings by Abdallah et al (12). It is not surprising because neonatal sepsis accounts for about 1 in 5 neonatal deaths in Uganda (25) and it is a significant contributor to neonatal deaths in most low and middle-income countries(26). Studies have shown that low birth weight is a risk factor for neonatal sepsis(27–29). Disease progression is fast, and deaths occur rapidly. Indeed, our study found that most of the deaths occurred at home before parents had taken action to save their infants. These findings are similar to what was found in Malawi (17). In part, this could reflect inadequacies by parents to recognize danger signs in neonates and seek care promptly as has been documented in Southwestern Uganda(30). On the other hand, this may be explained by the fact that caregivers who think an infant is likely to die may be less likely to seek urgent care, especially if the infant is fragile and small(31).

Predictors of mortality in this study were discharge weight <1200g, 5-minute Apgar score <7, and a diagnosis of neonatal sepsis during admission. Low birth weight neonates with discharge weight <1200g were 23 times more likely to die compared to heavier neonates. These results are comparable to what Abdallah et al, and Blencowe et al found (12,17); that the risk of post-discharge mortality increases with decreasing weight at discharge. This could be because smaller neonates, especially those with weight <1500g are more likely to be preterm(3), and have not yet attained oromotor maturity and appropriate homeostatic control. These neonates are therefore at greater risk of complications like hypothermia, hypoglycemia, and aspiration. They are also more likely to have immature immune systems (29). To compound this, they are likely to have been born before the maximal transplacental transfer of antibodies from the mother, which occurs in the last 8 weeks of gestation (32). They are therefore at increased risk of infections, hence mortality.

Low birth weight neonates with an Apgar score <7 at 5 minutes were 4 times more likely to die compared to those with an Apgar score ≥7. These results are in keeping with what Abdullah et al found in Bangladesh, what a systematic review by Ehrestein found, and what Cnattingius et al found in Sweden; that an Apgar score of <7 at 5 minutes was associated with an increased risk of mortality among LBW neonates (33–35). This may be due to complications of perinatal asphyxia like intraventricular hemorrhage, acute kidney injury, and hypoxic ischaemic encephalopathy. The association between prematurity and low Apgar score (36) may also explain the increased risk of mortality among these neonates.

Neonates who had sepsis during admission were 8 times more likely to die after discharge compared to those without this diagnosis. Neonatal sepsis is one of the major causes of neonatal mortality worldwide (37). The increased likelihood of post-discharge mortality among LBW neonates treated for neonatal sepsis during admission could reflect the persistence of some modifiable and non-modifiable risk factors of neonatal sepsis, like poor caregiver hygiene practices and prematurity. These neonates may therefore be at risk of re-infection and death even after discharge.

Improving survival of LBW neonates especially those with Apgar score <7 at 5 minutes, discharge weight <1200g, and /or have had sepsis during hospitalization requires intensive follow-up and care. There is a need to amend the guidelines for the discharge of LBW neonates so that they are discharged with weights >1200. Caregiver education on feeding, prevention of neonatal sepsis, early recognition of danger signs in neonates, and prompt care-seeking should be intensified. Studies aimed at prevention of post-discharge neonatal sepsis should be considered.

## Strengths and limitations of the study

This was the first study in Uganda to assess mortality and its predictors among LBW neonates following discharge from the SCBU. The prospective cohort study design used for this study allowed us to establish a temporal association between mortality and its predictors. The study was carried out at a tertiary care facility that receives many referrals. This may have caused selection bias. We relied on clinical symptoms identified and reported by caregivers to ascertain the possible causes of death.

## Conclusion

Neonatal mortality among low birth weight infants discharged from the Special Care Baby Unit at Mulago National Referral Hospital is high. Discharge weight of <1200g may be considered unsafe. Caregiver education about neonatal danger signs, and measures to prevent sepsis, aspiration, and cot death should be emphasized before discharge and during follow-up visits.

## Data Availability

All data are available upon request to the authors

https://data.mendeley.com/drafts/75rjpsfy96

## Abbreviations

ANC: Antenatal care
BPD: Bronchopulmonary dysplasia
CPAP: Continuous Positive Airway Pressure
ELBW: Extremely low birth weight
HIV: Human Immunodeficiency Virus
IMR: Infant Mortality Rate
IVH: Intraventricular hemorrhage
KMC: Kangaroo Mother Care
LBW: Low birth weight
LMIC: Low and middle-income countries
LNMP: Last Normal Menstruation Period
NEC: Necrotising enterocolitis
NICU: Neonatal Intensive Care Unit
NMR: Neonatal Mortality Rate
MNRH: Mulago National Referral Hospital
SCBU: Special Care Baby Unit
SDGs: Sustainable Development Goals
SGA: Small for Gestational Age
SSA: Sub-Saharan Africa
VLBW: Very low birth weight

## Acknowledgments

The authors are grateful to all the research assistants) and the study participants. We are also grateful to the staff at the Special Care Baby Unit, at Mulago National Referral Hospital.

## Author contributions

All authors made a substantial contribution to the work, whether that was in the conception, study design, implementation, data collection, analysis, and interpretation, or all these areas: manuscript drafting, review, and revision. All authors read and approved the final manuscript.

## Funding

This research did not receive any specific grant from funding agencies in the public, commercial, or not-for-profit sectors.

## Availability of data and materials

Available on request

## Declarations

Consent for publication

Not applicable.

## Competing interests

The authors declare that they have no competing interests.

---

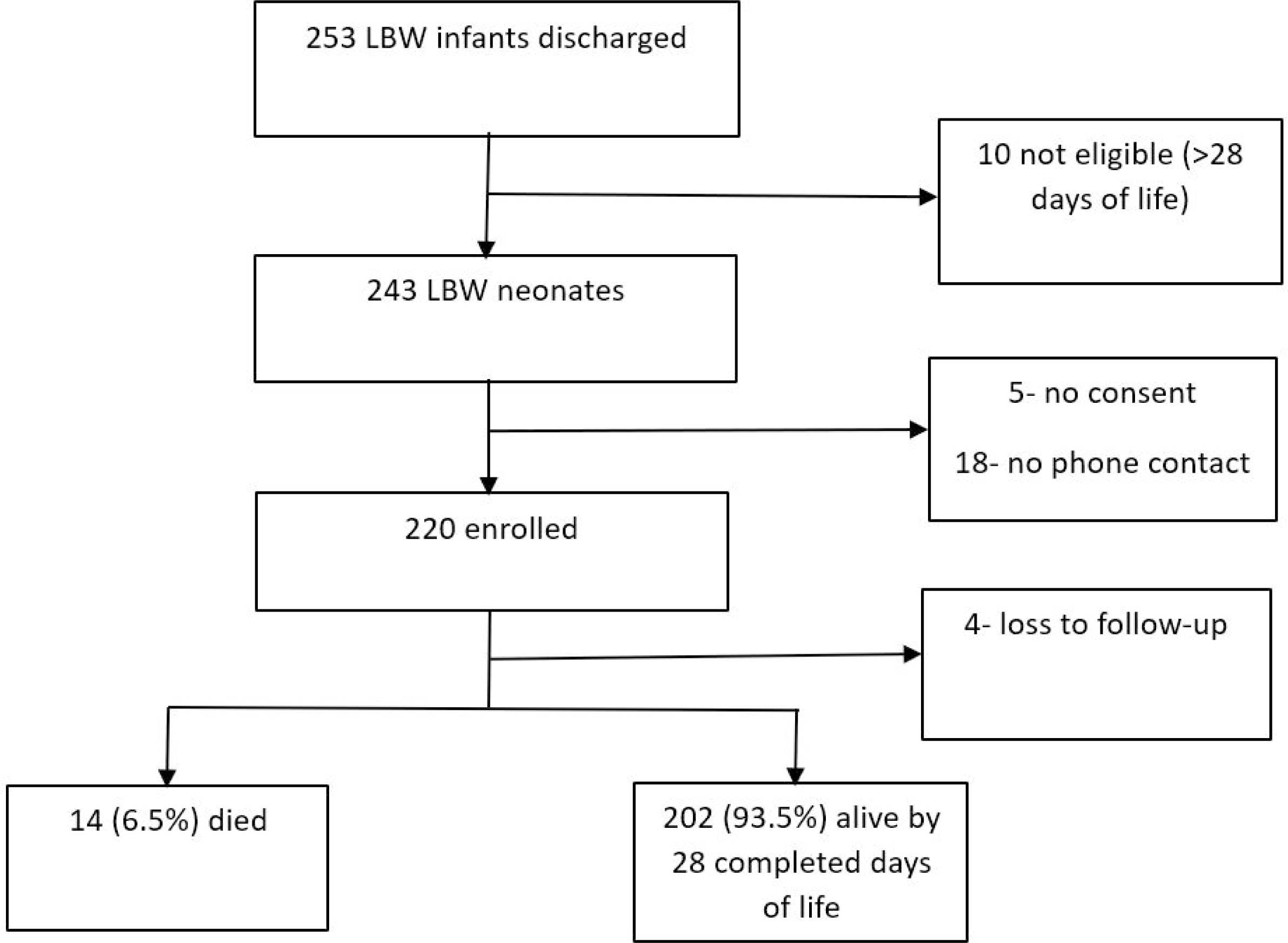

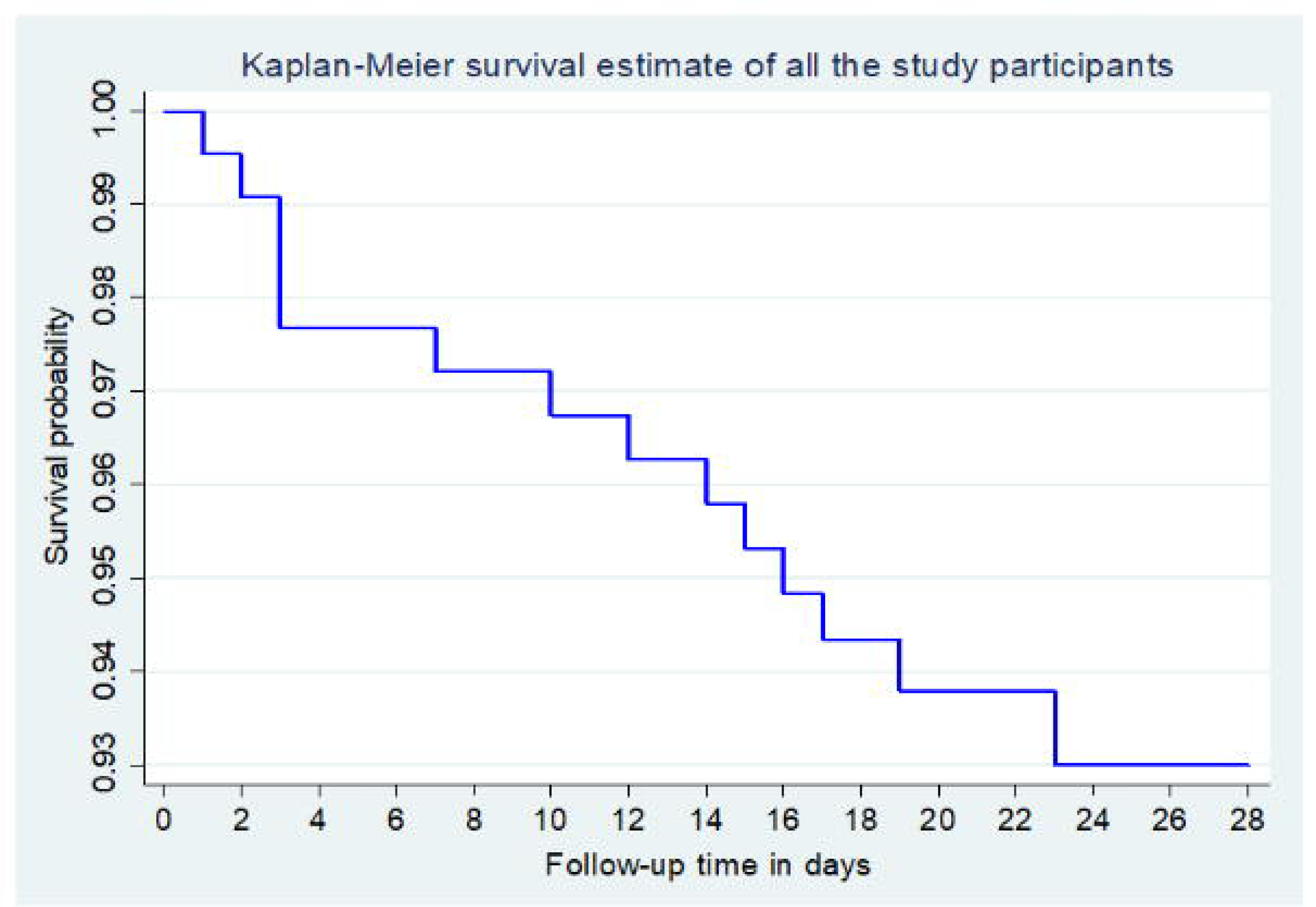

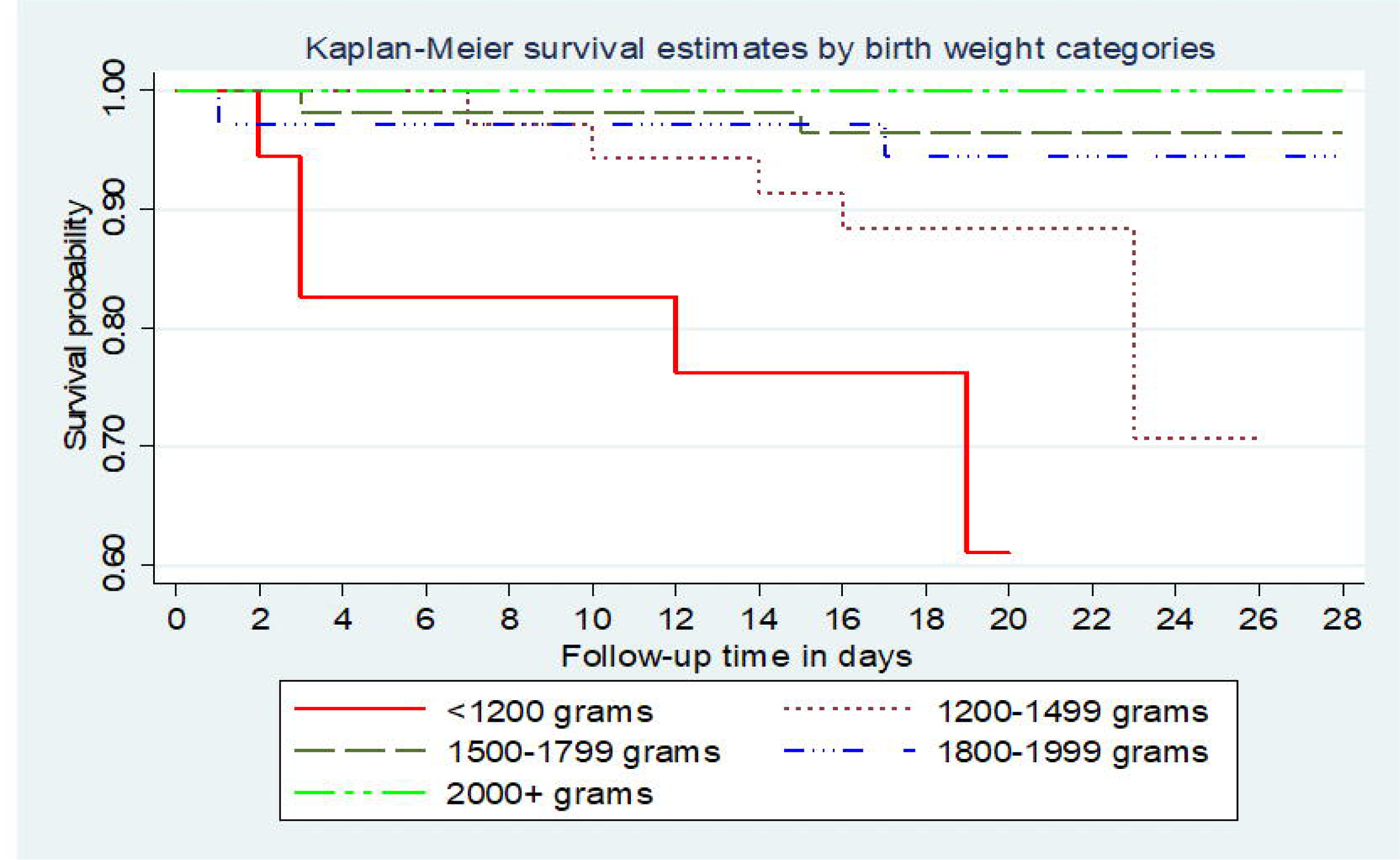

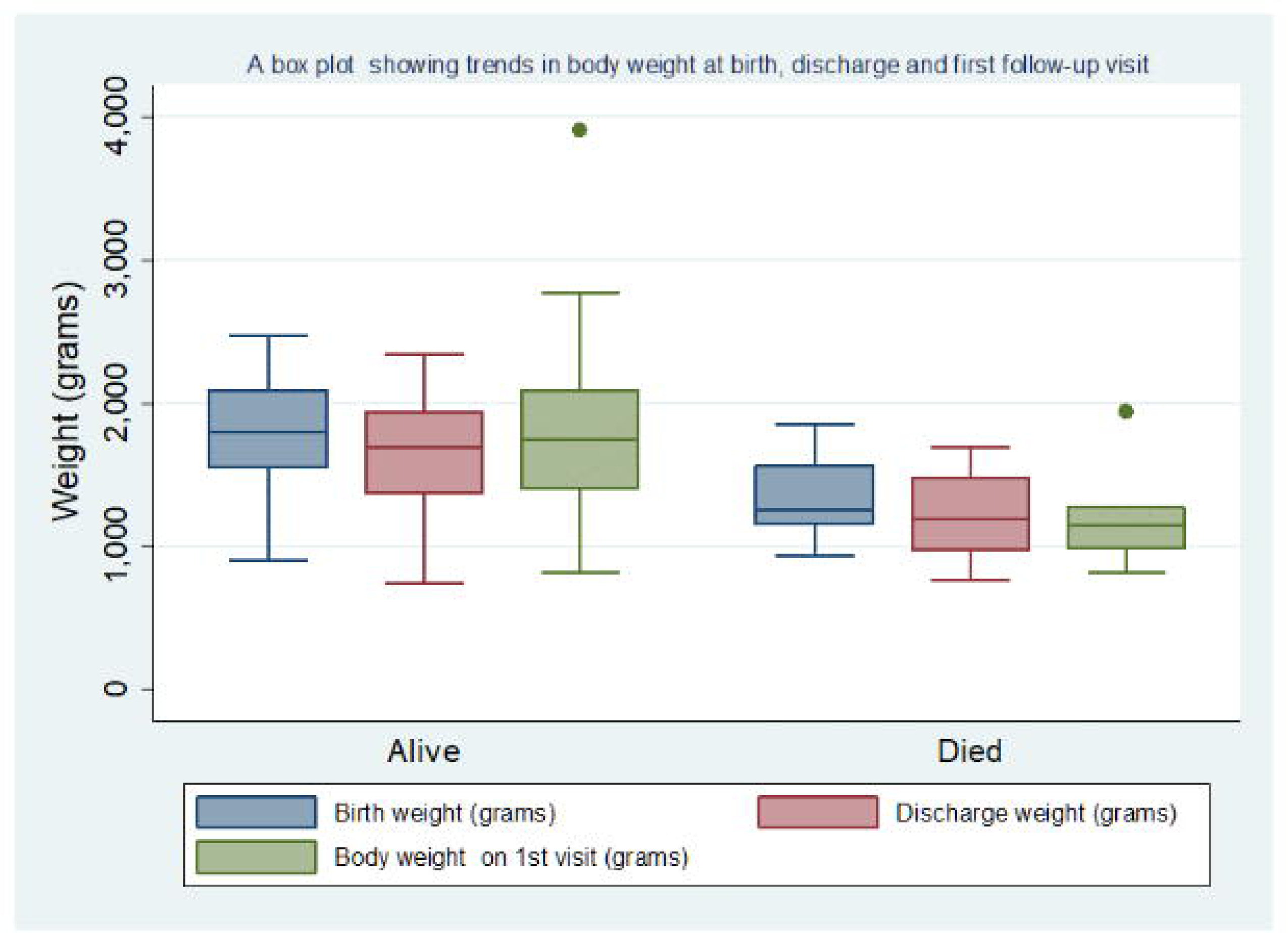

## Notes

### Competing Interest Statement

The authors have declared no competing interest.

### Clinical Protocols

https://data.mendeley.com/drafts/75rjpsfy96

### Funding Statement

This study did not receive any funding

### Author Declarations

The Makerere University School of Medicine Research Ethics Committee gave ethical approval for this work.

### Summary of Updates

The section on results has been updated so that Table 1 shows the baseline characteristics of study participants, and Table 2 shows the results from bivariate analysis. The conclusion in the main text has been revised to match the conclusion in the abstract.

